# Venous Thromboembolism and Intracranial Hemorrhage in Patients with High-grade Glioma

**DOI:** 10.1101/2020.01.17.20017475

**Authors:** Clara Borges, Carlota Lemos, Pedro Soares, Roberto Silva, Catarina Fernandes, Andreia Costa, Cláudia Caeiro, Cristina Sarmento

**Affiliations:** Medical Oncology department, Centro Hospitalar Universitário São João E.P.E., Porto, Portugal; Mathematics department, Polytechnic Institute of Viseu, Viseu, Portugal; Radio-oncology department, Centro Hospitalar Universitário São João E.P.E., Porto, Portugal; Anatomic pathology department, Centro Hospitalar Universitário São João E.P.E., Porto, Portugal

## Abstract

**Background:** Patients with high-grade glioma (HGG) are at increased risk of venous thromboembolism (VTE), throughout the course of disease. Prophylactic anticoagulation is not established among patients with HGG, outside of perioperative context, due to potential for intracranial hemorrhage (ICH) and the limited data available for predictive VTE scores in primary brain tumors. The aim of this study was to characterize VTE prevalence in adult patients with HGG and assess ICH risk during therapeutic anticoagulation.

**Methods:** Retrospective analysis of patients diagnosed with HGG at our institution, between 2009 and 2018. All adult patients proposed to systemic treatment were included into this study. Exclusion criteria was anticoagulation previous to diagnosis. VTE was defined as any radiographic-confirmed thrombus in the venous system. Risk factors of interest for VTE and bleeding risk scores were analyzed by chi-squared test and multivariate logistic regression. Survival analysis was performed using Kaplan-Meier method.

**Results:** A total of 410 patients were included of whom 31 (7,8%) developed a VTE, including 22 deep vein, 6 pulmonary and 3 central venous thrombosis. Twenty-nine patients with VTE had a WHO grade 4 glioma and 2 patients had grade 3 (anaplastic astrocytoma and oligodendroglioma).

In 22 cases, the VTE occurred during systemic treatment, more frequently during Temozolomide (n=15), followed by Irinotecan+Bevacizumab (n=6), Lomustine+Bevacizumab (n=1) and PCV (n=1). The median time between diagnosis and VTE was 10,11 months (95CI 6,46-14,79). Khorana score, age, ECOG performance status, smoking and obesity did not significantly differ in the VTE population. All VTE were initially treated with low molecular weight heparin (LMWH), of which 64.5% maintained LMWH, and the remainder switched to warfarin (19.4%) or to direct oral anticoagulant (16.1%). Six patients (19,4%) had spontaneous ICH under anticoagulation. Patients with grade 3 glioma (p=0,032) had significantly higher rates of ICH than grade 4. Patients with higher ECOG had significantly higher risk of ICH (OR 3,23 (95CI 1,18-8,81), p=0,022). HAS-BLED and ACCP bleeding scores were not associated with ICH. There was no significant difference in overall survival for TVE or ICH.

**Conclusion:** According to our data, ICH occurred in nearly 20% anticoagulated patients with HGG, as described in literature, and did not correlate with poorer prognosis. High ECOG performance status was an independent risk factor for ICH. Further effort towards better prediction models for VTE and ICH in HGG is warranted.

## Introduction

High-grade glioma (HGG) patients are at increased risk of venous thromboembolism (VTE), with incidence as high as 7,5-39% throughout the course of disease. The main concern when prescribing anticoagulation in HGG patients is the risk of intracranial hemorrhage (ICH).

Our study aims to characterize VTE risk and assess anticoagulation safety in HGG patients.

## Methodology

Retrospective analysis of adult patients with a diagnosis of World Health Organization grade III or IV glioma, who were offered systemic treatment at a tertiary referral hospital, between 2009 and 2018. Previous anticoagulation was an exclusion criteria.

The primary endpoint was VTE, defined as radiographic-confirmed thrombus in venous system. The secondary endpoint was ICH risk during anticoagulation.

Analysis of risk factors for VTE and ICH was performed by chi-squared test and multivariate logistic regression; and overall survival (OS) analysis by Kaplan-Meier.

The trial had hospital ethics comitee approval and patients’ informed consent.

## Results

There was a total of 410 adult patients with HGG included in the study. Clinical and pathological characteristics are described in Table 1. Thirty-one patients developed a VTE (7,8%), including 22 deep vein thrombosis, 6 pulmonary embolisms and 3 central venous thrombosis.

**Table 1.**
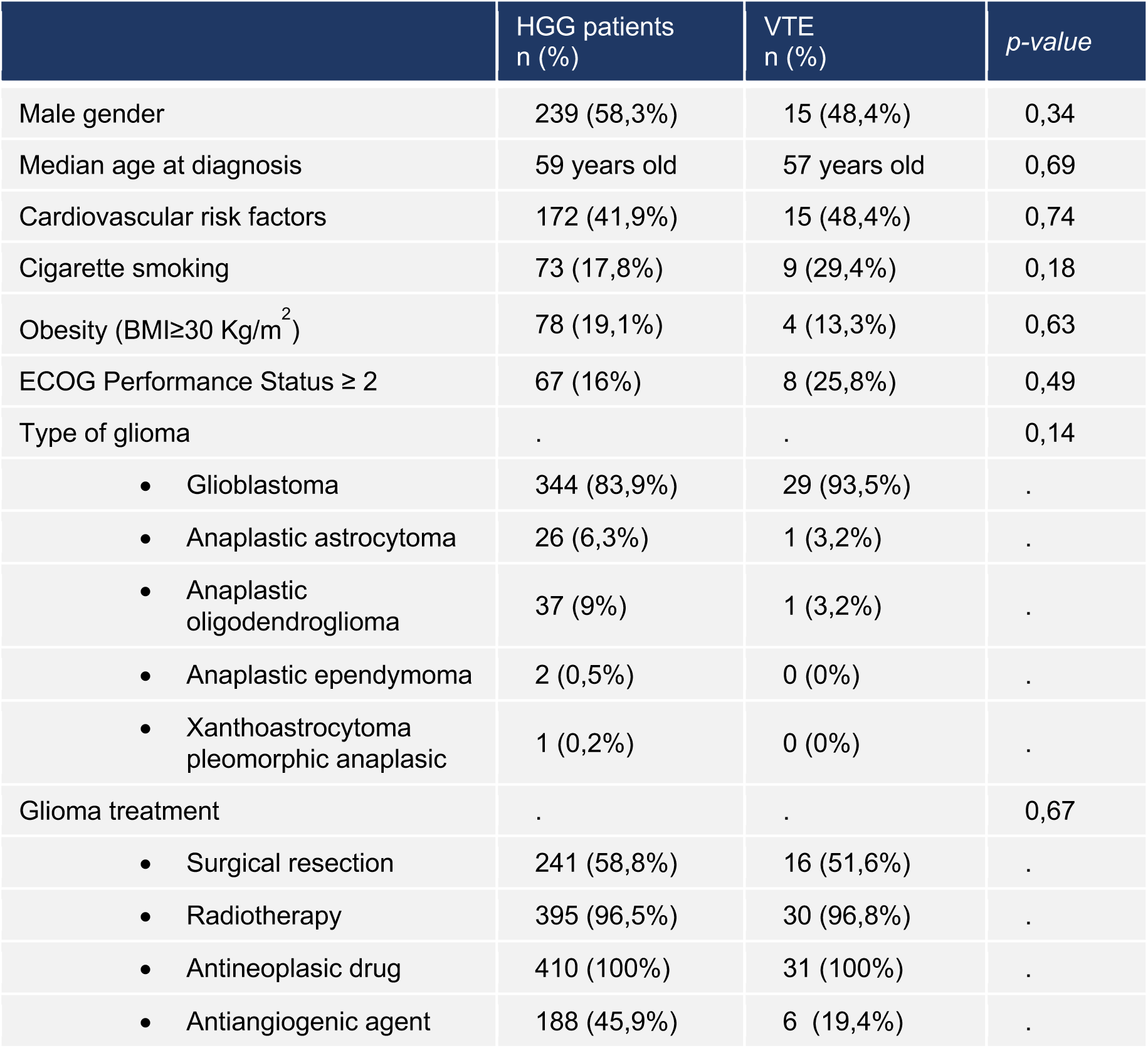
Patient characteristics.

|  | HGG patients<br>n (%) | VTE<br>n (%) | *p-value* |
| --- | --- | --- | --- |
| Male gender | 239 (58,3%) | 15 (48,4%) | 0,34 |
| Median age at diagnosis | 59 years old | 57 years old | 0,69 |
| Cardiovascular risk factors | 172 (41,9%) | 15 (48,4%) | 0,74 |
| Cigarette smoking | 73 (17,8%) | 9 (29,4%) | 0,18 |
| Obesity (BMI≥30 Kg/m<sup>2</sup>) | 78 (19,1%) | 4 (13,3%) | 0,63 |
| ECOG Performance Status ≥ 2 | 67 (16%) | 8 (25,8%) | 0,49 |
| Type of glioma | . | . | 0,14 |
| • Glioblastoma | 344 (83,9%) | 29 (93,5%) | . |
| • Anaplastic astrocytoma | 26 (6,3%) | 1 (3,2%) | . |
| • Anaplastic oligodendroglioma | 37 (9%) | 1 (3,2%) | . |
| • Anaplastic ependymoma | 2 (0,5%) | 0 (0%) | . |
| • Xanthoastrocytoma pleomorphic anaplastic | 1 (0,2%) | 0 (0%) | . |
| Glioma treatment | . | . | 0,67 |
| • Surgical resection | 241 (58,8%) | 16 (51,6%) | . |
| • Radiotherapy | 395 (96,5%) | 30 (96,8%) | . |
| • Antineoplastic drug | 410 (100%) | 31 (100%) | . |
| • Antiangiogenic agent | 188 (45,9%) | 6 (19,4%) | . |

Older age, worse performance status, smoking habits, obesity or other cardiovascular risk factors were not significantly associated with VTE.

The median time from diagnosis to VTE was 10,11 months (95CI 6,46-14,79).

Most patients had a VTE during systemic treatment (71%): 15 patients (68,2%) on Temozolomide; 5 (16,1%) on Irinotecan + Bevacizumab; 1 (3,2%) on Lomustine + Bevacizumab; 1 (3,2%) on PCV protocol. Three patients had VTE on post-operatory setting (9,7%). Six patients were on best supportive care (19,4%).

All VTE patients were initially treated with low molecular weight heparin (LMWH). Most patients maintained long-term anticoagulation with LMWH (n=20; 64,5%), 6 patients were treated with warfarin (19,4%) and 5 with direct oral anticoagulant (16,1%).

Khorana score for VTE risk was calculated as shown in Table 2 and 3. There were no high risk patients and most patients with VTE were low risk (64,5%).

**Table 2.** Khorana score for VTE risk in cancer patients.

| Variable | Score |
| --- | --- |
| Very high-risk tumor (stomach, pâncreas) | 2 |
| High-risk tumor (lung, gynecologic, genitourinary excluding prostate) | 1 |
| Hemoglobin level <10 g/L or use of growth factors | 1 |
| Prechemotherapy leukocyte count $\geq 11 \times 10^9$ /L | 1 |
| Prechemotherapy platelet count $\geq 350 \times 10^9$ /L | 1 |
| Body mass index $\geq 35$ Kg/m<sup>2</sup> | 1 |

**Table 3.** VTE risk.

|  | HGG patients<br>n (%) | VTE<br>n (%) | *p-value* |
| --- | --- | --- | --- |
| Khorana Score | . | . | 0,24 |
| • High risk (Score $\geq 3$) | 0 | 0 | |
| • Intermediate risk (Score 1-2) | 180 (45,8%) | 11 (35,5%) |  |
| • Low risk (Score < 1) | 213 (54,2%) | 20 (64,5%) |  |

There was not a significant association between higher Khorana score and VTE incidence (p=0,24).

Six patients (19,4%) had spontaneous ICH under anticoagulation. There were no other major bleed bleedings.

Three (50%) patients were anticoagulated on LMWH. The other half was treated with warfarin.

The median time from VTE to ICH was 2,88 months (95CI 0,46-5,30).

Risk factor analysis for ICH suggested that ICH was significantly more frequent in patients with worse performance status (ECOG PS ≥2 42,8% vs ECOG PS <2 0%). Multivariate logistic regression confirmed that higher ECOG PS was significantly associated with higher ICH risk (OR 3,23 (95 CI 1,18-8,81) p=0,022)

There was a significantly higher rate of ICH observed among patients with grade III glioma than grade IV (100% vs 13,8%, p=0,032).

We calculated three bleeding scores to our anticoagulated population – HAS-BLED score (Hypertension, abnormal liver or renal function, previous stroke, prior history or predisposition to bleeding, labile or unstable INR, age, drugs and alcohol), PANWARDS score (platelet count, albumin, congestive heart failure, age, race, diastolic blood pressure and previous stroke or transient ischemic attack) and ACCP score (age, previous bleeding, cancer, metastatic cancer, renal failure, liver failure, thrombocytopenia, previous stroke, diabetes, anemia, antiplatelet therapy, poor anticoagulant control, comorbidity and reduced functional capacity, recent surgery, frequent falls, alcohol abuse and nonsteroidal anti-inflammatory drug).

Neither was associated with higher ICH rate in our population. According to HAS-BLED and PANWARDS risk score, all patients were low risk. According to ACCP bleeding risk score, there were 21 high risk (score > 1) anticoagulated patients (67,7%). However, high risk was not associated with higher ICH rate when compared to low/intermediate risk (p=0,534).

There was no significant difference in OS for patients with VTE (17,87 vs 14,43 months, p=0,944).

There was also no significant difference in OS for ICH (20,20 vs 17,43 months, p=0,412), as shown in Figure 1

**Figure 1.**
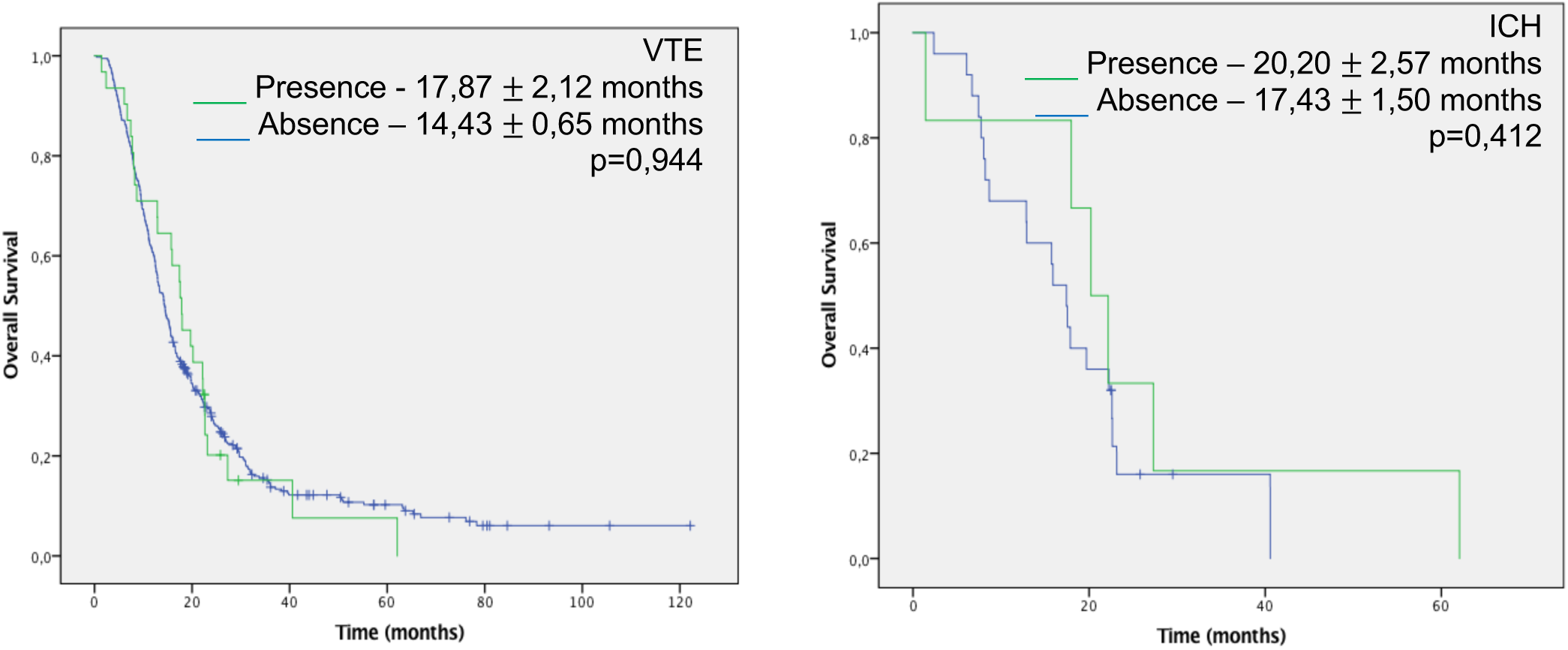
Kaplan-Meier survival curves for OS, according to VTE and ICH.

## Discussion

VTE is a frequent occurrence in HGG patients. The etiology of thrombosis is multifactorial, including a latent hypercoagulable state, neurosurgical procedures and immobility due to neurologic deficits that further predisposes patients to VTE. Khorana score is not validated for predicting VTE risk in HGG population and further effort towards better prediction models is warranted.

Anticoagulation should be carefully considered for HGG patients with a VTE event, as these patients have a high bleeding risk. According to our data, ICH occurred in nearly 20% anticoagulated patients and was more frequent among grade III glioma patients (p=0,032). ECOG performance status was an independent risk factor for ICH (OR 3,23 (95 CI 1,18-8,81) p=0,022).

Additional studies identifying which HGG patients are at highest risk of ICH are of paramount importance to facilitate the development of a bleeding risk score that guides physicians’ decisions.

## Data Availability

The data that support the findings of this study are available from the corresponding author, upon reasonable request.

## References

• Mantia C, Uhlmann EJ, et al. Predicting the higher rate of intracranial hemorrhage in glioma patients receiving therapeutic enoxaparin. Blood 2017;129(25):3379–85

• Taillibert S, Tailandier L, et al. Venous thrombosis in patients with high-grade glioma. Curr Opin Oncol 2015;27:516–21

• Perry, JR. Thromboembolic disease in patients with high-grade glioma. Neuro-Oncology 2012;14:73–80

• Perry JR, Julian JA, et al. PRODIGE: a randomized placebo-controlled trial of dalteparin low-molecular-weight heparin thromboprophylaxis in patients with newly diagnosed malignant glioma. J Thromb Haemost 2010;8:1959–65

• Perry SL, Bohlin C, et al. Tinzaparin prophylaxis against venous thromboembolic complications in brain tumor patients. J Neurooncol 2009;95(1):129–34

